# Six Latent Variables Underlie MDS-UPDRS Scores and Reveal a Dissociation Between Patient- and Clinician-Assessed Parkinson’s Disease Symptoms

**DOI:** 10.64898/2026.08.10.26360084

**Authors:** Bhadra Santhi Kumar, Mark Humphries

**Affiliations:** University of Nottingham

## Abstract

The Movement Disorder Society’s Unified Parkinson’s Disease Rating Scale (MDS-UPDRS) is the global standard for characterising Parkinson’s Disease (PD) in clinical contexts. However, the specific symptom phenotypes it captures remain poorly understood, potentially limiting its value for diagnosis, prognosis, and stratifying patients. To address this, we developed a spectral estimation approach to find the unique latent variables captured by the 60 scores of MDS-UPDRS parts I, II, and III from 852 sporadic PD patients. Our analysis revealed six latent variables that robustly captured variation between patients and generalised across cohorts. The primary variable encoded symptom laterality, while others encoded distinct clinical features including tremor severity, and revealed an unexpected dissociation between patient self-reported symptoms and clinician-assessed symptoms, highlighting potential gaps in how PD is currently evaluated and understood. Our findings open the door to precise MDS-UPDRS phenotyping of patients for treatments and clinical trials.

## 1 Introduction

Parkinson’s disease (PD) is considered the fastest growing neurological disorder[1, 2]. It impacts the motor and cognitive abilities of patients and can severely impact their quality of life[3, 4]. Like most neurodegenerative diseases, PD cannot be cured but its symptoms can be managed effectively, improving the patient’s quality of life. Treatment plans primarily aim to improve motor symptoms by medically substituting dopamine with its precursors. In advanced PD cases, surgical interventions like Deep Brain Stimulation (DBS) can significantly improve patients’ quality of life.

The Movement Disorder Society’s Unified Parkinson’s Disease Rating Scale (MDS-UPDRS) is consistently used world wide to assess PD symptoms[5, 6, 7]. The MDS-UPDRS is a set of questions evaluating the severity of a patients’ motor and non motor symptoms. MDS-UPDRS parts I and II are the patient’s self-reported evaluation of their motor and cognitive abilities and their quality of life. MDS-UPDRS part III is the clinician’s evaluation of the patient’s motor symptoms. MDS-UPDRS is a critical tool in PD since it is the basis for deciding treatment plans, prognosis, and even eligibility for DBS implantation [8, 5, 6, 7].

Despite its widespread use, it remains unclear what variation across patients is captured by the MDS-UPDRS. Two patients with identical MDS-UPDRS scores can have widely differing collections of symptoms, implying differing forms of PD with differing trajectories and prognosis. Identifying the latent variables in MDS-UPDRS would reveal the range of patient phenotypes it captures. By capturing these phenotypes, such latent variables could offer significant value in predicting the course of the disease and in supporting the stratification of patients for personalised treatment and clinical trials. This raises a crucial question: what are the unique latent variables represented in the MDS-UPDRS?

Several studies have looked at the latent structure of MDS-UPDRS and its predecessor the UPDRS using factor analysis techniques [9, 10, 11, 12, 13, 14]. These studies agreed that the MDS/UPDRS contains a handful of latent variables, but disagreed on how many and what they were, limiting the ability to generalise the latent structure across cohorts.

To overcome these limitations, we sought to objectively identify the latent variables captured by the MDS UPDRS assessment. We developed a spectral estimation approach[15], which statistically distinguishes genuine latent variables from chance correlations between MDS-UPDRS scores, providing a data-derived upper bound on the latent variables in the MDS-UPDRS. Applying this to MDS-UPDRS data from 852 patients in the Parkinson’s Progression Marker’s Initiative (PPMI) cohort, we find MDS-UPDRS has a latent structure of six unique symptom variables. The structure was robust within and across cohorts, stable across disease progression, and could accurately reconstruct held-out patient data. This latent structure can capture the heterogeneity of PD using MDS-UPDRS assessments, offering a basis for improved diagnosis, prognosis, and stratification of patients for trials.

## 2 Results

### 2.1 MDS-UPDRS data has a six dimensional structure beyond noise

To determine the latent symptom variables captured by MDS-UPDRS, we considered parts I-III of the baseline assessment of 852 sporadic PD patients when they were enrolled in the PPMI cohort. Parts I-III of the MDS-UPDRS are defined using 60 features scored on a severity scale of 0 (normal) to 4 (severe). As these patients were not on any dopamine-replacement medication at the time of their baseline assessment, part IV, which measures the motor complications arising from long term medication, was omitted.

We found the correlation matrix of MDS-UPDRS Parts I-III scores had a block diagonal structure for the 852 patient cohort, consistent with a low-dimensional structure of MDS-UPDRS (Fig. 1a). To qualitatively confirm this, we applied Principal Component Analysis (PCA) to the data, which revealed the first few components explained a considerable fraction of the variance in MDS-UPDRS parts I-III scores across patients (Fig.1b).

**Figure 1:**
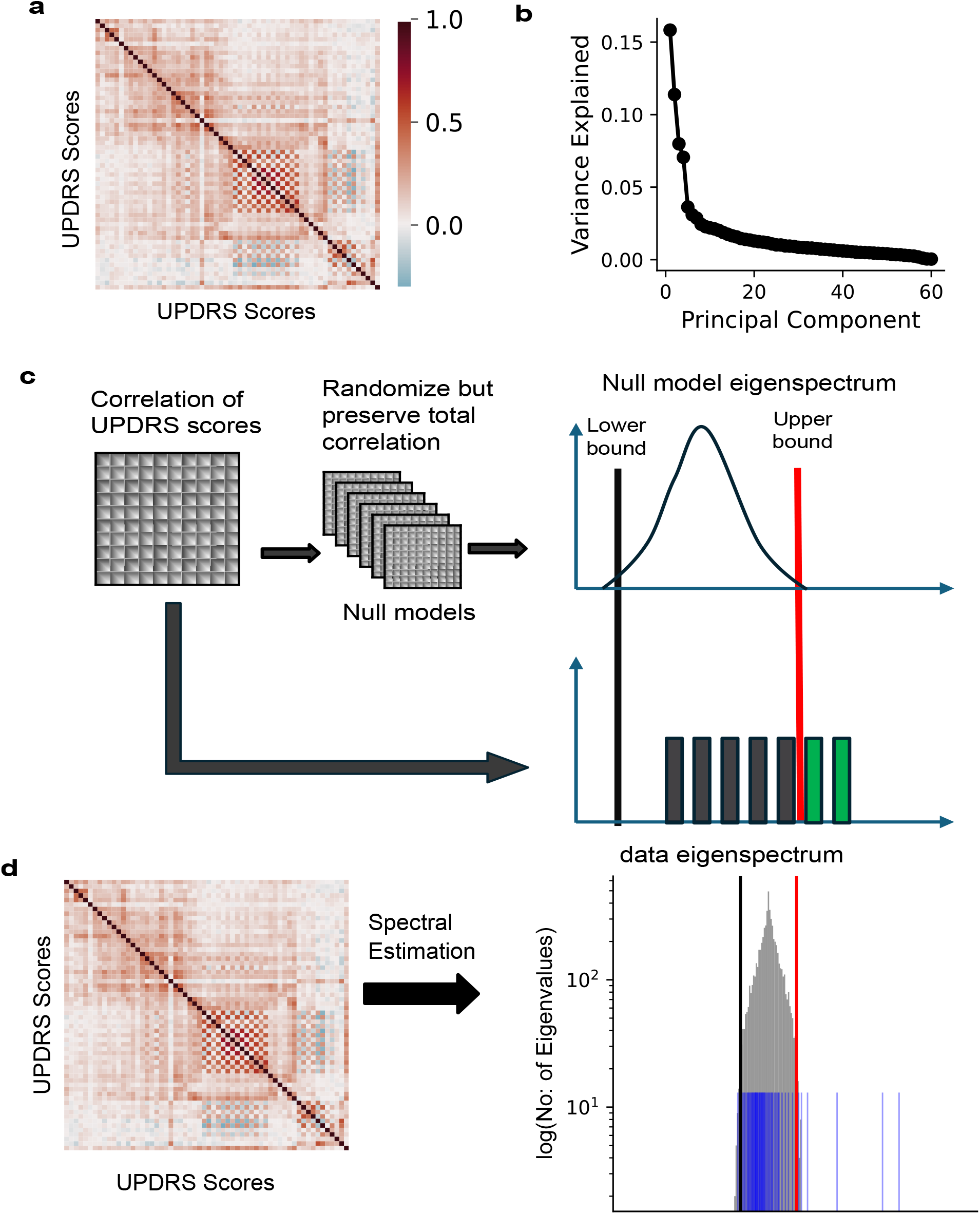
The MDS-UPDRS parts I-III have a low dimensional structure. (a) Correlation between MDS-UPDRS scores of N=852 patients in the PPMI cohort. (b) Proportion of variance explained by each principal component of the correlation matrix in panel a. (c) Schematic of spectral estimation. The red vertical line denotes the upper bound of the eigenvalues of a data correlation matrix predicted by the null model. Data eigenvalues (bars) exceeding the upper bound (green bars) correspond to latent dimensions of the data. (d) Spectral estimation of the latent structure of MDS-UPDRS scores. The grey histogram shows the eigenvalue distribution of the null model, with its lower and upper bounds marked by the black and red lines, respectively. The blue bars beyond the upper bound represent excess eigenvalues of the MDS-UPDRS correlation matrix (panel a) compared to the null model, indicating a six-dimensional structure in the MDS-UPDRS data.

To objectively determine the low-dimensional structure, we developed a spectral estimation approach [15]. Spectral estimation identifies a set of data dimensions that depart from a null model of high-dimensional data. It does this by generating a set of null model correlation matrices that preserve the total correlation of each variable while randomizing the specific pairwise relationships (Fig.1c; Methods). These null models thus represent the correlation structure contributed by noise, including rater noise, a known issue in MDS-UPDRS scoring [16]. By comparing if and how the data correlation departs from these null models, we isolate a unique set of data dimensions that reflect the latent structure of the data beyond what could be explained by noise. By using a specified null model, this entirely unsupervised approach thus offers a more objective identification of the size and content of MDS-UPDRS’ latent structure.

Applying spectral estimation to the correlation matrix of the MDS-UPDRS data, we found six dimensions that exceeded the upper bound of the null distribution (Fig.1d). MDS-UPDRS parts I-III thus have a six-dimensional latent structure that exceeds the correlation structure predicted by noise. We call the subspace defined by these six dimensions the “baseline” space. Each of its dimensions identifies a latent variable captured by MDS-UPDRS.

### 2.2 Dimensions of the baseline space are clinically interpretable

We sought to interpret the symptoms captured by each dimension of the baseline space. Figure 2 plots the loading on each of dimension for each of the 60 features of MDS-UPDRS parts I-III. High loadings of the same sign on the same dimension indicate persistently co-occurring clinical features; high loadings of opposite sign indicate persistently anti-correlated clinical features; low loadings indicate little contribution to this dimension. The color coding groups MDS-UPDRS features associated with major symptom categories of bradykinesia, rigidity, tremor, and axial symptoms in the MDS-UPDRS III, and the motor and non-motor symptoms in MDS UPDRS I-II. The acronyms used for the MDS-UPDRS scores are explained in Table 1.

**Table 1:** Mapping of MDS-UPDRS items to symptom domains and their clinical descriptions.

| Symptom domain | UPDRS code | Clinical description |
| --- | --- | --- |
| Cognitive | NP1COG<br>NP1HALL<br>NP1DPRS<br>NP1ANXS<br>NP1APAT<br>NP1DDS | Cognitive impairment<br>Hallucinations<br>Depression<br>Anxiety<br>Apathy<br>Dopamine dysregulation syndrome |
| Motor | NP1PAIN<br>NP1URIN<br>NP1CNST<br>NP1LTHD<br>NP2SPCH<br>NP2SALV<br>NP2WALK<br>NP2EAT<br>NP2TURN<br>NP2TRMR<br>NP2RISE<br>NP2FREZ<br>NP2HWRT<br>NP2SWAL | Pain<br>Urinary problems<br>Constipation<br>Lethargy<br>Speech<br>Saliva<br>Walking<br>Eating<br>Turning in bed<br>Tremor<br>Arising from chair<br>Freezing of gait<br>Handwriting<br>Swallowing |
| Other | NP1SLPN<br>NP1SLPD<br>NP1FATG<br>NP2DRES<br>NP2HYGN<br>NP2HOBB | Sleep problems - night<br>Sleep problems - day<br>Fatigue<br>Dressing<br>Hygiene<br>Hobbies |
| Bradykinesia | NP3FTAPR<br>NP3FTAPL<br>NP3HMOVR<br>NP3HMOVL<br>NP3PRSPR<br>NP3PRSPL<br>NP3TTAPR<br>NP3TTAPL<br>NP3LGAGR<br>NP3LGAGL | Finger tapping (right)<br>Finger tapping (left)<br>Hand movements (right)<br>Hand movements (left)<br>Pronation-supination of hand (right)<br>Pronation-supination of hand (left)<br>Toe tapping (right)<br>Toe tapping (left)<br>Leg agility (right)<br>Leg agility (left) |
| Rigidity | NP3RIGRU<br>NP3RIGLU<br>NP3RIGRL<br>NP3RIGLL<br>NP3RIGN | Rigidity (right upper limb)<br>Rigidity (left upper limb)<br>Rigidity (right lower limb)<br>Rigidity (left lower limb)<br>Rigidity (neck) |
| Tremor | NP3PTRMR<br>NP3PTRML<br>NP3KTRMR<br>NP3KTRML<br>NP3RTARU<br>NP3RTALU<br>NP3RTARL<br>NP3RTALL<br>NP3RTALJ<br>NP3RTCON | Postural tremor (right hand)<br>Postural tremor (left hand)<br>Kinetic tremor (right hand)<br>Kinetic tremor (left hand)<br>Rest tremor amplitude (right upper limb)<br>Rest tremor amplitude (left upper limb)<br>Rest tremor amplitude (right lower limb)<br>Rest tremor amplitude (left lower limb)<br>Rest tremor amplitude (jaw)<br>Constancy of rest tremor |
| Axial symptoms | NP3SPCH<br>NP3FACXP<br>NP3RISNG<br>NP3GAIT<br>NP3FRZGT<br>NP3POSTR<br>NP3PSTBL | Speech<br>Facial expression<br>Arising from chair<br>Gait<br>Freezing of gait<br>Posture<br>Postural stability |

**Figure 2:**
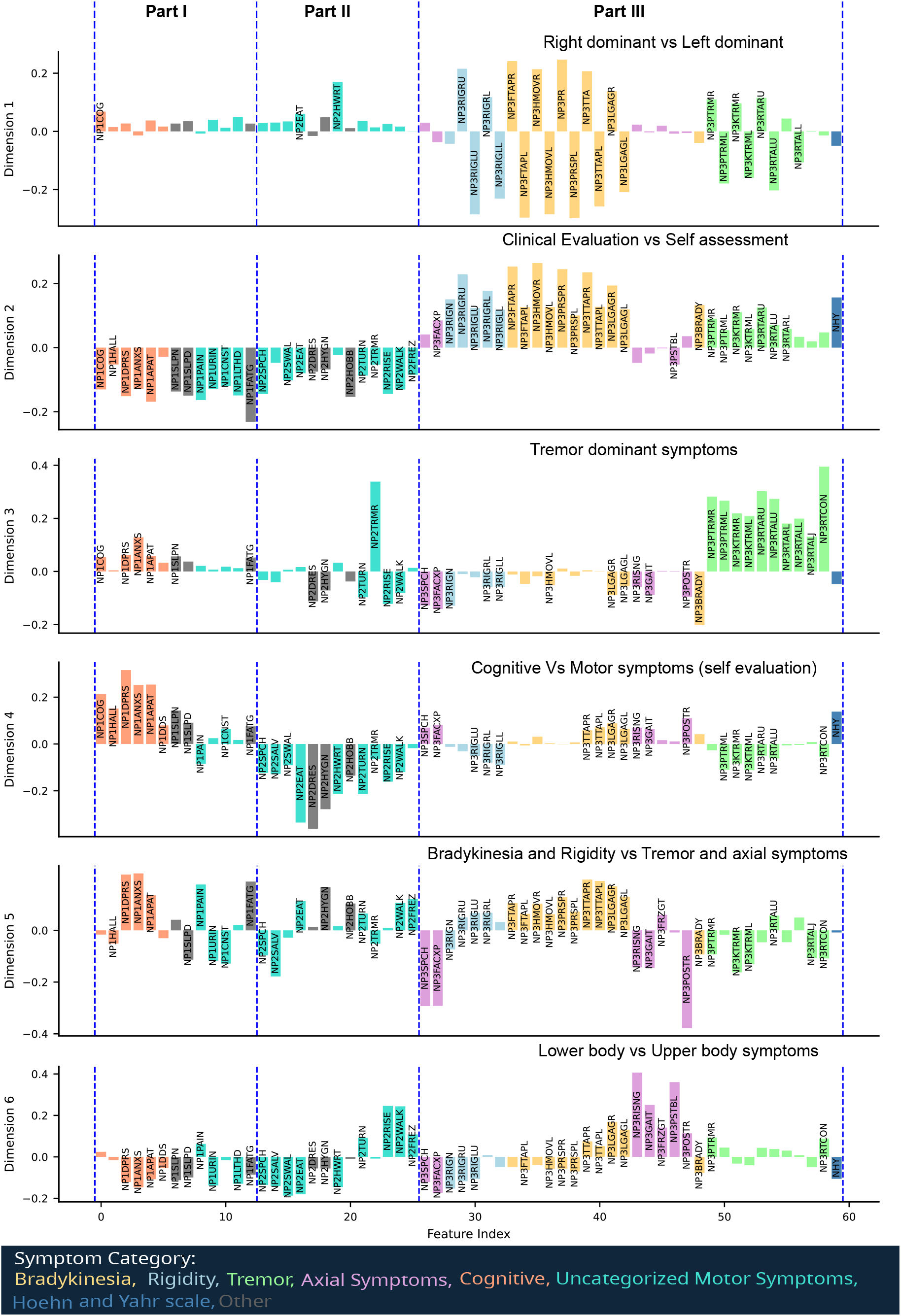
Interpretation of the baseline space. We plot the loadings of all MDS-UPDRS parts I, II, and III features on each dimension. Colors represent assignments of MDS-UPDRS features to a symptom category (Methods). Blue dashed lines separate MDS-UPDRS parts I, II and III. Dimensions are assigned latent variable names based on the domi5nant features they capture – see text. Dimensions are ordered top-to-bottom by their decreasing explanatory power, as determined by their distance above the null model’s upper bound.

We found the first dimension differentiated whether patients’ symptoms were dominant on the left or right side of the body (Fig. 2): Loadings for left-side symptoms were systematically of opposite sign to those of right-side symptoms. The second dimension clearly differentiated patients’ self-evaluation of symptoms (parts I and II of MDS-UPDRS) from clinical assessment of motor symptoms (part III). As parts I and II capture many aspects of cognition and movement that affect quality of life, this dimension suggests little relationship between perceived quality of life and the externally-judged severity of motor symptoms. The third dimension captured co-occurring features of tremor. It also showed they were anti-correlated with other motor symptoms. This dimension thus differentiated patients with tremor-dominant PD from other forms. The fourth dimension showed that, within the self-assessment of symptoms (MDS-UPDRS I,II), cognitive features co-occurred, as do motor features. But it also showed that cognitive and motor features have opposite signs of loadings. This dimension thus differentiated patients that self-reported strong cognitive symptoms from those that reported strong motor impacts on everyday life. The fifth dimension differentiated patients with co-occurring bradykinesia and rigidity from those with co-occurring tremor and axial symptoms. The sixth dimension differentiated patients with upper and lower body symptoms. Thus, our unsupervised approach not only shows MDS-UPDRS has six latent variables, but also revealed those latent variables to be clinically interpretable.

### 2.3 The baseline space is robust

The number of latent variables in the MDS-UPDRS and what they represent could depend on the size of the patient cohort used to discover them. To check if this latent structure was stable, we generated further baseline spaces using increasingly large subsets of the available baseline data from the sporadic PD cohort.

The dimensionality of the subspaces generated by these subsets stabilized beyond 400 patients (Fig.3a). This suggests the latent structure of MDS-UPDRS has a consistent size. To test if the latent symptom variables represented by the dimensions were also consistent, we compared the baseline space for each subset of patients to the reference baseline space from the full cohort of 852 sporadic patients. We found the subset-derived and the reference baseline space were highly similar across all subsets (Fig.3b), and so had highly similar representations of the MDS-UPDRS data on each dimension. Notably, even subsets containing less than half of the full cohort maintained strong alignment with the reference baseline space. The baseline space and the latent variables it captured were thus robust to the number of patients used to construct it, suggesting we had identified a consistent latent structure within MDS-UPDRS Parts I-III.

**Figure 3:**
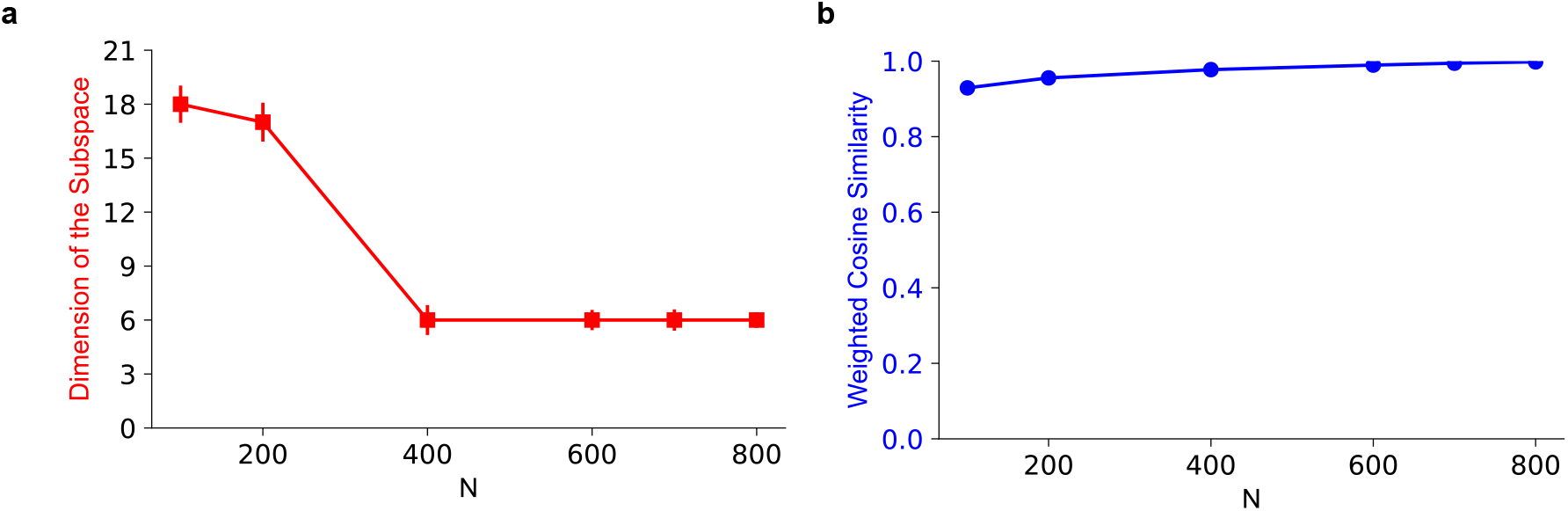
The baseline space of MDS-UPDRS is robust. (a) The number of dimensions found by spectral estimation on subsets of *N* patients from the sporadic PD cohort. Error bars plot medians and standard error of the median [17] over 50 samples of each patient subset. (b) Similarity of the discovered subspaces. We plot the weighted cosine similarity between the six principal dimensions of the reference baseline space and each subspace built from a subset of cohort data. A similarity of 1 indicates the reference baseline space and the subspace were identical; a similarity of 0 indicates the two were orthogonal. Error bars plot mean and standard error over the same patient subsets as panel a: the standard error was typically around 0.001, hence not visible in the plot.

### 2.4 The baseline space generalises to unseen patient data

This consistency of the baseline space of MDS-UPDRS implies that it should generalise well to new patient data. We tested this hypothesis in three ways.

First, we split the full sporadic cohort (N=852) into an 80% training set to estimate a baseline space and a held-out 20% testing set to assess that space’s ability to generalise to new patients. We assessed generalisation by how well we could reconstruct all 60 MDS-UPDRS scores of the held-out patients from the projection of their MDS-UPDRS data into the baseline space created from the training set of patients. To establish lower bounds for reconstruction quality, we also projected the held-out patient’s data into random subspaces, generated by independently permuting the loading values within each dimension – this ensured that the overall distribution of values in each dimension was preserved while removing any relationships between dimensions. Reconstruction quality was measured by root mean squared error (RMSE), to measure reconstruction fidelity, and the Pearson correlation coefficient, to assess preservation of linear relationships.

We found that reconstructing MDS-UPDRS scores of held-out patients using the baseline space was significantly better than the lower bound, in both their linear correlation with the true data (*N* = 250, *t* = 133, *p* < 0.001)(Fig. 4a) and their RMSE (*N* = 250, *t* = −66, *p* < 0.001)(Fig. 4b). Notably, the quality of reconstruction of the MDS-UPDRS scores from held-out patients was as good as the reconstruction of the scores from patients in the training set (Supplementary Fig. S1 a,b). Together, these results suggest the baseline space generalises well to new patients.

**Figure 4:**
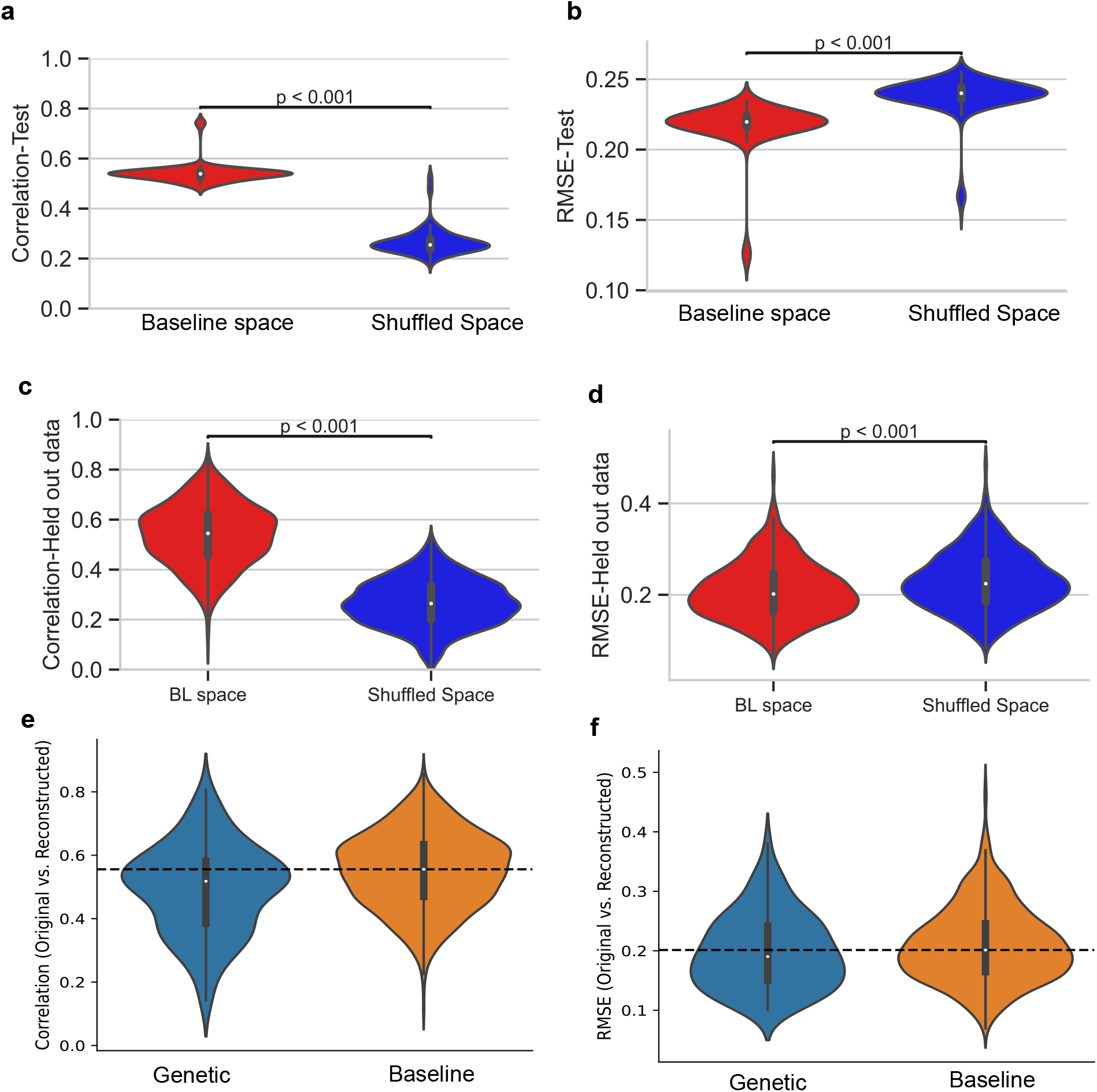
The baseline space generalises to unseen patient data. (a, b) Pearson correlation and Root Mean Square Error (RMSE) between the test cohort’s true and reconstructed MDS-UPDRS scores. Distributions are shown over 250 estimates from 50 repeats of 5-fold cross validation. (c, d) Pearson correlation and RMSE between the held-out patient’s true and reconstructed MDS-UPDRS data. (e,f) Reconstruction of genetic cohort patient data (N=90) from the sporadic cohort’s (N=852) baseline space. The dashed line shows the median of the baseline cohort’s reconstruction metrics. Distributions are over the patients in each cohort. P-values in panels a-d are from two-sided paired t-tests.

Second, we modelled a clinical scenario in which a new patient is assessed using MDS-UPDRS and their data projected into the baseline space. To do so, we held-out each patient in turn from the N=852 sporadic cohort, constructed the baseline space from the N=851 patients, and assessed our ability to reconstruct the held-out patient’s MDS-UPDRS scores. We found reconstruction of the held-out patient was significantly better than the lower bound in both the linear correlation with the true data (*N* = 852, *t* = 53.0, *p* < 0.001)(Fig. 4c) and their RMSE (*N* = 852, *t* = −41, *p* < 0.001)(Fig. 4d). Furthermore, the quality of reconstruction for held-out patients was comparable to that obtained for patients in the training set (Supplementary Fig. S1c,d), demonstrating that the baseline space would generalise well in a clinical scenario.

Third, we tested the ability of the baseline space to generalise across cohorts. The PPMI database also contained a “genetic” cohort of N=90 PD patients with an identified risk variant in LRRK2, GBA or SNCA genes. We projected the MDS-UPDRS scores from the initial assessment of this genetic cohort on to the baseline space of the sporadic cohort, and reconstructed the genetic cohort’s scores. The reconstruction of the genetic cohort’s MDS-UPDRS scores (correlation median value=0.52, RMSE median value=0.2) was about as good as the reconstruction of the sporadic cohort’s own scores (correlation median value=0.56, RMSE median value=0.2) (Fig. 4e,f). Consequently, the baseline space generalises well not only to unseen sporadic patient data, but also to an entirely new cohort of patients.

### 2.5 The baseline space of MDS-UPDRS is stable across disease progression

The six-dimensional latent structure of MDS-UPDRS so far described was derived from the initial assessment of patients in the PPMI cohort at or near their diagnosis. With time, PD symptoms worsen and patients start medication to control them. This raises the question of whether the latent variables captured at baseline would generalise across the worsening of symptoms with time and the effect of medication. To address this, we used the baseline space to reconstruct each patient’s full set of 60 MDS-UPDRS scores taken at each later assessment – a “visit” – in the PPMI cohort. As patients started dopamine replacement therapy after their initial assessment, their later visits include clinician’s assessment of UPDRS part III both ON and OFF medication. We assessed reconstruction of both sets of scores.

The median correlations between the original and reconstructed scores where high and stable across visits for both ON- and OFF-medication states (Fig.5). Correlations ranged approximately from 0.41 to 0.68 ON medication and 0.49 to 0.60 OFF medication, encompassing the baseline’s median correlation of 0.56. These results indicate the latent structure of PD symptoms captured by MDS-UPDRS is stable over disease progression and medication status.

**Figure 5:**
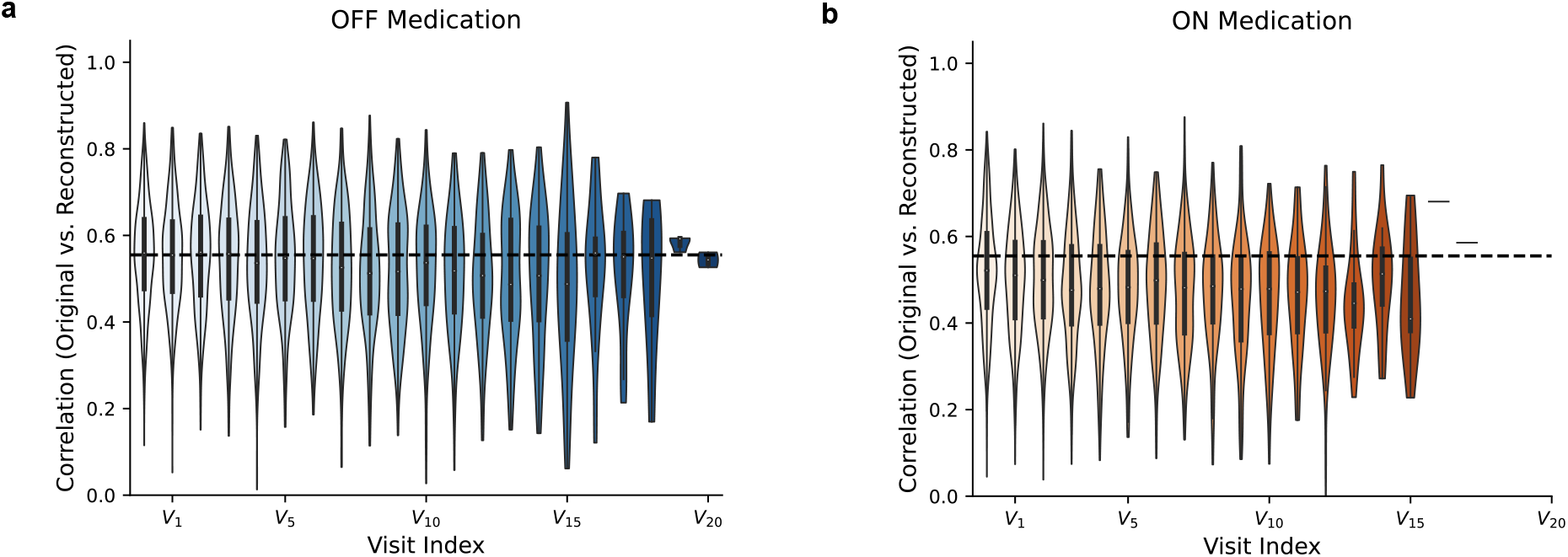
Stability of the baseline space across disease progression. (a) The correlation between true and reconstructed OFF medication MDS-UPDRS scores at each assessment. Scores were reconstructed by projecting MDS-UPDRS data at each visit on to the baseline space. Dashed line is the median correlation between true and reconstructed baseline MDS-UPDRS scores using that space. (b) As for panel a, for ON medication MDS-UPDRS assessments.

### 2.6 Quantifying disease progression using the latent variables of MDS-UPDRS

The above analyses had established that the latent structure of MDS-UPDRS has clinically interpretable latent variables and is stable over disease progression. We next asked if the latent structure could give insights into the progression of specific symptoms and how that progression is affected by medication.

We first quantified patients’ symptom load at their initial assessment. We projected the patients’ baseline MDS-UPDRS data onto each of the six symptom dimensions. This gave us a ‘burden score’ for each patient on each latent variable: a burden score of zero indicated the patient had no clinical features captured by that variable, a higher burden score indicated more severe clinical features, and the sign of the burden score indicated which co-occurring clinical features within that variable the patient had more of. We measured the variance of the burden score across the cohort to capture the distribution of severity for each latent symptom variable (Fig. 6a).

**Figure 6:**
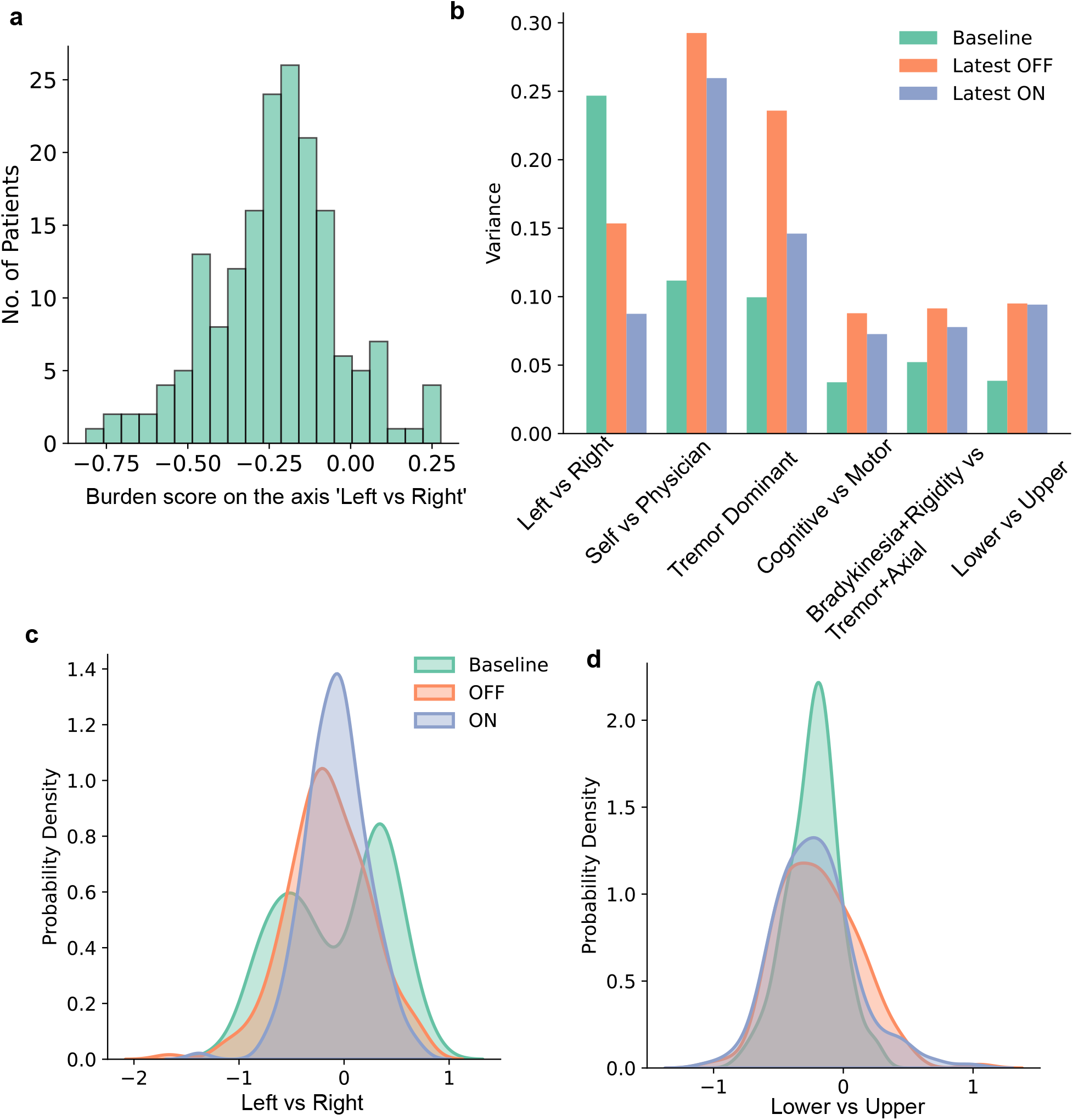
The latent structure of disease progression and medication effects in MDS-UPDRS. (a) The cohort’s distribution of burden scores at baseline for the left vs right symptom latent variable. (b) Variance of patients’ burden scores on each of the six latent variables for their baseline assessment and their latest assessments OFF and ON medication. (c) Kernel density estimate of the distribution of burden scores on the Left vs Right latent variable at baseline and ON and OFF medication states. (d) Kernel density estimate of the distribution of burden scores on the Lower vs Upper latent variable at baseline and ON and OFF medication states

Symptom progression on each latent variable was tracked by comparing the variance of burden scores at baseline and at the patients’ most recent assessment OFF medication. Figure 6b shows that the variance of burden scores increased between the baseline (blue bars) and latest OFF medication assessment (red bars) for five of the six latent variables. These increases in variance indicate increases in symptom severity over time and are in line with expected symptom progression: for example, the variance of the cohort’s burden score on the tremor dominant latent variable more than doubled, consistent with increasingly strong tremor over time from tremor-dominant patients.

The exception to the increase in variance over time was the latent variable that captured the laterality of symptoms: the variance of the cohort’s burden score decreased for this variable from baseline to the most recent assessment. We interpret this decrease as capturing the spread of symptoms from the initially dominant body side to the other. Indeed, we found that the initial bimodal distribution of the burden score on this laterality variable at baseline changed to a narrower, unimodal distribution by the most recent assessment OFF medication (Fig. 6c).

The effect of medication on each latent symptom variable was tracked by comparing the variance of the burden score ON and OFF medication at the latest assessment. Figure 6b shows that medication reduced the variance of symptom burden in five of the six latent variables (compare orange and cyan bars). Again, this is in line with the expected effects of medication in ameliorating symptoms. The exception was the variable differentiating upper and lower body symptoms, on which medication had minimal impact (Fig 6d).

## 3 Discussion

The MDS-UPDRS is the standard instrument for assessing the symptom burden of Parkinson’s patients. But what symptom variations within Parkinson’s disease are captured by the MDS-UPDRS’ 60 scored features is unclear. We used an objective approach to show that MDS-UPDRS data has a six dimensional latent structure. This low dimensional space could not only accurately and robustly represent the baseline assessment data from which it was derived, but generalised across cohorts, progression stages, and medication states.

Previous studies of the dimensionality of MDS-UPDRS and its predecessor, the UPDRS [18], used factor analysis. They consistently found a low dimensional structure in their cohort data [10, 11, 12, 14, 19, 13]. However, a key limitation of traditional factor analysis lies in the determination of the number of factors, which is predominantly based on some subjective criterion, often visual inspection of a plot showing the variance explained by each dimension (a scree plot, similar to Fig. 1b). This procedure introduces subjectivity and renders the results dependent on the specific dataset under investigation, constraining the generalisability of the findings. In contrast, spectral estimation derives the dimensionality in a data-driven way by testing whether the observed structure differs from the latent structure expected under a null model. This makes the identified dimensions more robust, reproducible, and less dependent on the dataset.

We found each of the six dimensions was a clinically-relevant latent variable within MDS-UPDRS. The first and sixth latent variables distinguish symptoms based on body halves, the first differentiates patients with left or right-hand side dominant and the sixth differentiates patients with lower and upper body symptoms. These latent variables, though found unsupervised, nonetheless capture the well known laterality of PD motor symptoms [20, 21] and the upper versus lower limb split in some symptoms [22, 23].

The second latent variable distinguishes clinician-evaluated symptoms from self-evaluated symptoms. This indicates that, while the scores are correlated within the self evaluation (MDS-UPDRS I and II) and within the clinician’s evaluation (MDS-UPDRS III), there is no correlation between the two sets of evaluations. Prior studies have found similar discrepancies between the self evaluation and clinician’s assessment in PD patients[24, 25, 26].

The third latent variable captured the well-established distinction between the tremor-dominant and other phenotypes of PD [27]. This is also consistent with the tremor-related variables previously identified within UPDRS data using factor analysis [13] and item response theory [28]. Our findings contribute further evidence that tremor behaves as a distinct dimension of motor impairment, whereas other symptoms – such as bradykinesia, rigidity, and axial impairment – do not demonstrate the same degree of independence, supporting the clinical utility of distinguishing tremor-dominant and non-tremor phenotypes of PD.

The fourth latent variable revealed a clear cognitive–motor structure within the self-assessment of Parts I and II: cognitive and neuropsychiatric items from Part I loaded with the same sign on this dimension, whereas motor-related items from Part II loaded with the opposite sign. This separation is consistent with prior work showing that several Part I items map onto objective cognitive performance across multiple domains [29], as well as with the established organisation of the MDS-UPDRS, in which Part I captures non-motor (including cognitive and neuropsychiatric) experiences and Part II reflects motor experiences of daily living [6]. It also implies patients can be differentiated by their dominant expression of either non-motor or daily living issues.

The fifth latent variable differentiated patients with bradykinesia and rigidity from those with tremor- and axial-dominated features. Prior factor-analytic studies of UPDRS and MDS-UPDRS Part III typically identify bradykinesia, rigidity, tremor, and axial symptoms as partially separable domains, with axial and gait-related items often clustering with bradykinesia and rigidity rather than with tremor [10, 11, 6, 30]. In contrast, our analysis of the full MDS-UPDRS, incorporating Parts I, II, and III, reveals a relative dissociation of bradykinesia and rigidity from axial symptoms. This suggests that integrating self-reported measures of daily living and non-motor experiences may capture additional heterogeneity in symptom structure that is not apparent when considering motor examination scores alone.

The six dimensional latent structure of MDS-UPDRS has several implications for how PD is assessed and monitored. As we demonstrated, it provides a standardised space in which to quantify disease progression. Future work will assess if it is easier to track patient’s progression trajectories and distinguish different types of PD progression in this latent space. Similarly, the specific trajectory taken by a patient in this latent space could give insights into their disease progression and prognosis. Comparison of patients’ trajectories ON and OFF medication could provide additional information on the impact of medication on symptoms. Furthermore, the evidently crucial latent variable separating self evaluation from clinician assessment highlights that patients experience PD in ways that are not fully captured by clinical evaluation. This observation suggests the development of more patient-centered measures of treatment response.

The latent structure of MDS-UPDRS we describe opens the door to the precise phenotyping of patients. As we have shown (Figure 4), any patient’s MDS-UPDRS phenotype can be captured in six latent variables, by projecting that patient’s MDS-UPDRS assessment on to the six dimensions of the baseline space we describe here. This six-dimensional phenotype of each patient is well-suited to stratifying patients: it could be used to predict the outcome of established treatments, like Levodopa, for individuals; equally, it could be used in clinical trials to link patients’ phenotypes to the effectiveness of the trialled intervention.

## 4 Methods

### 4.1 Data preprocessing

We used the MDS-UPDRS scores collected in the Parkinson’s Progression Marker’s Initiative (PPMI) database. The sporadic cohort of the PPMI study had 889 patients at the time of access (April 2024). After removing the patients with missing scores, we had 852 patients with a complete set of MDS-UPDRS I, II and III assessments consisting of 60 scores measured during the baseline visit.

The test scores are ordinal numbers ranging from 0 to 4 indicating five levels of severity (normal, slight, mild, moderate and severe) of symptoms. We used rank normalization to normalize each score into the range [0,1]:

1. Use the ordinal value of a score to define its rank (1 to *R*). Eg; scores 0 − 4 will have a rank of 1 − 5
2. Normalize the scores (*x*) of each patient using the rank of the score (*r*) and the maximum rank of that score (*R*)

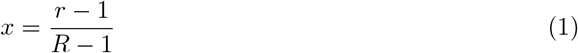

We defined the maximum rank *R* of the score from its MDS-UPDRS definition, rather than from the data itself, to ensure that the normalization remained consistent and robust across diverse datasets.

### 4.2 Dimensionality reduction

The rank normalized baseline data (852 × 60) was used for studying the structure of MDS-UPDRS. The PCA on the data was done using the scikit learn library https://scikit-learn.org/stable/. The spectral estimation was performed on the correlation matrix derived from the MDS-UPDRS data (60 × 60). Our spectral estimation algorithm is described in ref.[15]; a MATLAB toolbox is available at https://github.com/mdhumphries/NetworkNoiseRejection; we used the Python implementation of this algorithm by Thomas J Delaney used in ref [31] and available at (https://github.com/thomasjdelaney/Network_Noise_Rejection_Python)). We generated 100 null models to establish the expected lower and upper bounds on the eigenvalues of the MDS-UPDRS modularity matrix —-defined as the MDS-UPDRS correlation matrix minus the expected network derived from the null model —-in the absence of meaningful latent structure. The algorithm returned the *d* eigenvectors of the MDS-UPDRS modularity matrix, corresponding to the *d* eigenvalues that exceeded the null model’s predicted upper bound. We gather these in matrix *U* (60 × *d*): we refer to each eigenvector as a “dimension”, and *U* as the *d*-dimensional subspace.

#### 4.2.1 Analysis of loadings

To analyze the loadings along the six dimensions of the baseline space, we grouped the MDS-UPDRS part III scores into broader groups of bradykinesia, rigidity, tremor and axial symptoms following [32]. The scores for MDS-UPDRS I and II were grouped into motor and cognitive symptoms where this was clear from their description; we label the remaining Part I and II scores as “other”. The grouping of the symptoms is given in Table 1.

### 4.3 Weighted cosine similarity

To assess the robustness of the 6-dimensional baseline space *U*_*B*_ constructed from the full baseline dataset, we compared it to subspaces *U*_*N*_ obtained from random subsets of patients of size *N* ∈{100, 200, 400, 600, 700, 800}. For each subset, we computed a weighted cosine similarity between *U*_*B*_ and *U*_*N*_ based on the principal angles between these subspaces. Specifically, we first computed the inner product matrix

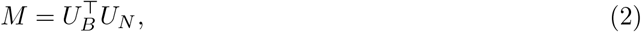

The singular values of *M*, obtained via SVD, are known to correspond directly to the cosines of the principal angles between the two subspaces:

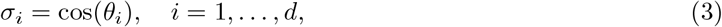

with *d* = min(dim(*U*_*B*_), dim(*U*_*N*_)). Rather than explicitly computing the angles *θ*_*i*_, we used the singular values {*σ*_*i*_} as subspace similarity measures.

To aggregate across dimensions, we computed a weighted cosine similarity:

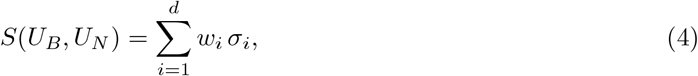

The weights *w*_*i*_ were derived from the eigenvalue spectrum of the full baseline subspace, normalized to sum to one:

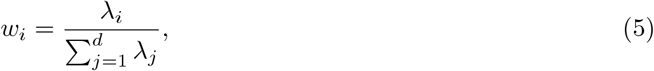

where *λ*_*i*_ denotes the eigenvalues associated with the eigenvectors in *U*_*B*_.

### 4.4 Validation of the low-dimensional representation

To test if the low-dimensional representation of MDS-UPDRS scores was robust, we tested the reconstruction of held-out patient data from the six-dimensional space.

For each held-out patient, we reconstructed their data as follows:

1. Project the 60-dimensional data vector **x** ∈ ℝ^60^ onto the 6-dimensional space spanned by eigenvectors 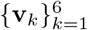

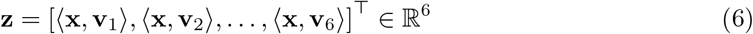
2. Reconstruct the original data from the 6-dimensional representation **z**:

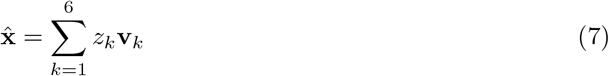
3. Measure the quality of reconstruction using two metrics:
  a. **Root Mean Square Error (RMSE)**: Let **x** ∈ℝ^*D*^ be the original data vector and 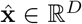 the reconstructed vector. The RMSE is computed as:

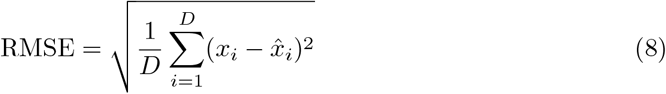
  b. **Pearson Correlation Coefficient**: The Pearson correlation coefficient between **x** and 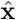 is given by:

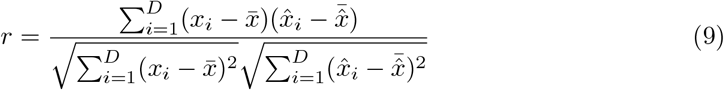
4. To evaluate whether the low-dimensional reconstruction is better than chance, we define and utilize a control space:
  a. Construct a random (control) version of the low-dimensional space by independently shuffling the values within each eigenvector:

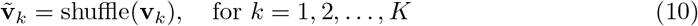

where **v**_*k*_ is the *k*-th eigenvector and 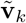 is its shuffled version. The shuffling is done element-wise within each vector, destroying meaningful structure while preserving the value distribution.
  b. Use the shuffled basis vectors 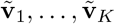 to project the original data and reconstruct it:

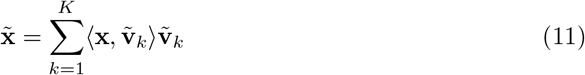
  c. Compute the reconstruction error (RMSE and Pearson correlation) for the shuffled reconstructions 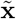, as above.

## Data Availability

Data used in the preparation of this article was obtained on [2024-04-30] from the Parkinson’s Progression Markers Initiative (PPMI) database (www.ppmi-info.orgaccess-data-specimens/download-data), RRID:SCR 006431. For up-to-date information on the study, visit www.ppmi-info.org.

## Code Availability

Code to reproduce all analyses in this paper is available at: https://github.com/Humphries-Lab/UPDRS_latent_structure_paper/.

## 5 Acknowledgements

This work was funded by Innovate UK grant [10036282]. We thank Tom Gilbertson for comments on a draft of this manuscript. PPMI – a public-private partnership – is funded by the Michael J. Fox Foundation for Parkinson’s Research and funding partners, including AbbVie, Alamar Biosciences, Aligning Science Across Parkinson’s (ASAP), Arrowhead Pharma, Arvinas, AskBio, BIAL, BioArctic, Biohaven, BlueRock Therapeutics, Bristol Myers Squibb, Calico Labs, Capsida Biotherapeutics, Critical Path Institute, DaCapo Brainscience, Denali, Edmond J. Safra Foundation, Eli Lilly, Gain Therapeutics, GE Healthcare, Genentech, GSK, Insitro, Johnson & Johnson Innovative Medicine, Lundbeck, Merck, Neumora, Neuron23, Novarti, Olink, Regeneron, Roche, Sanofi, Tenvie, UCB, Vanqua Bio, Voyager Therapeutics, The Weston Family Foundation.

## 6 Supplementary material

**Supplemental Figure S1:**
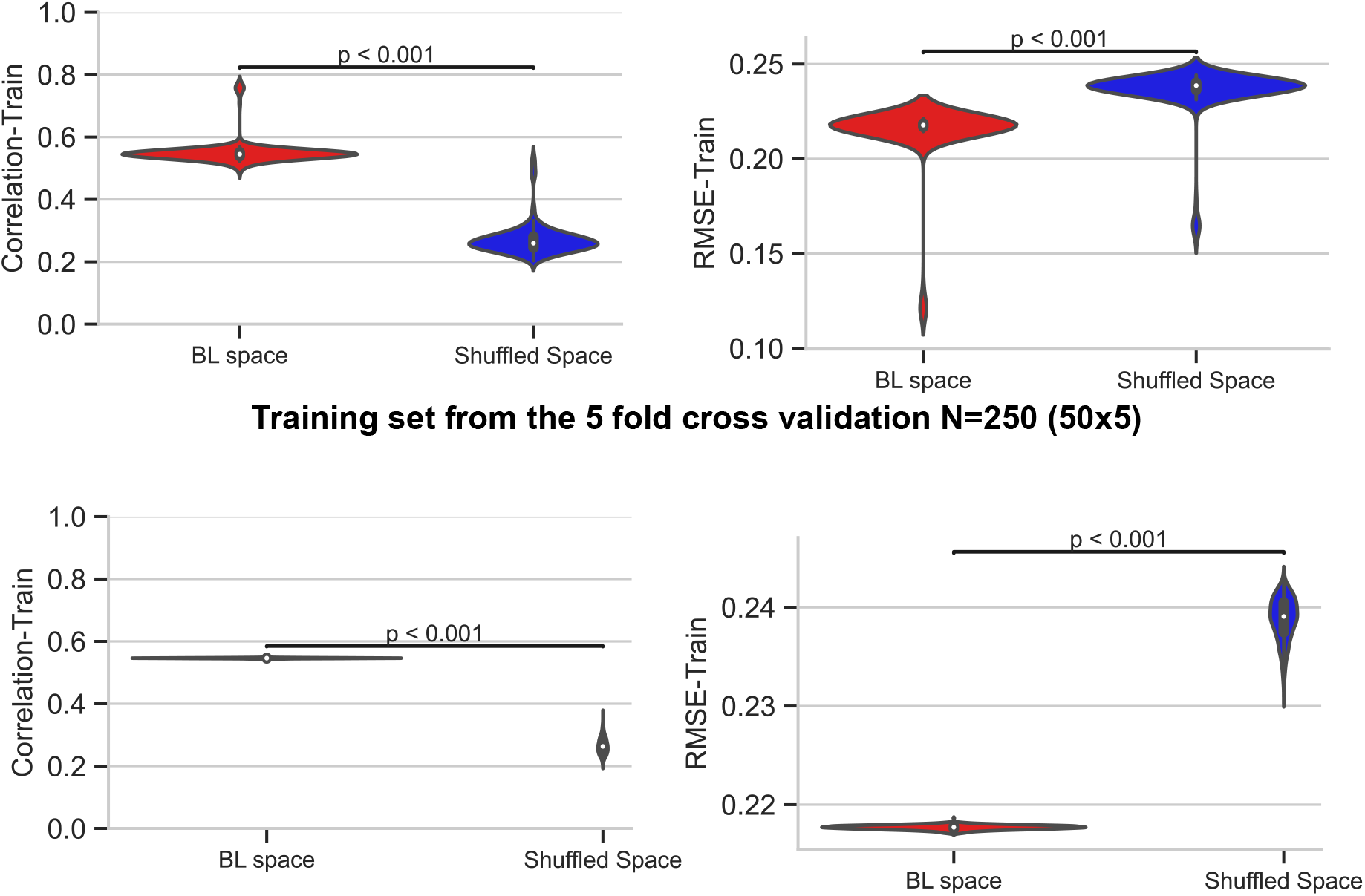
Reconstruction of training data MDS-UPDRS scores using the baseline subspace. (a, b) Pearson correlation and Root Mean Square Error (RMSE) between the train cohort’s true and reconstructed MDS-UPDRS scores for 5-fold cross-validation (N=250:50×5 cross validation). (c, d) As for panels a and b, for the training cohort for the held-out patient assessment (N=852).

## References

[1] E. R. Dorsey, T. Sherer, M. S. Okun, and B. R. Bloem, “The emerging evidence of the parkinson pandemic,” Journal of Parkinson’s disease, vol. 8, no. 1, pp. S3–S8, 2018.

[2] D. Su, Y. Cui, C. He, P. Yin, R. Bai, J. Zhu, J. S. T. Lam, J. Zhang, R. Yan, X. Zheng, J. Wu, D. Zhao, A. Wang, M. Zhou, and T. Feng, “Projections for prevalence of parkinson’s disease and its driving factors in 195 countries and territories to 2050: modelling study of global burden of disease study 2021,” BMJ, vol. 388, p. e080952, 2025.

[3] D. Aarsland, L. Batzu, G. M. Halliday, G. J. Geurtsen, C. Ballard, K. Ray Chaudhuri, and D. Weintraub, “Parkinson disease-associated cognitive impairment,” Nature reviews Disease primers, vol. 7, no. 1, p. 47, 2021.

[4] N. Zhao, Y. Yang, L. Zhang, Q. Zhang, L. Balbuena, G. S. Ungvari, Y.-F. Zang, and Y.-T. Xiang, “Quality of life in parkinson’s disease: A systematic review and meta-analysis of comparative studies,” CNS neuroscience & therapeutics, vol. 27, no. 3, pp. 270–279, 2021.

[5] C. G. Goetz, S. Fahn, P. Martinez-Martin, W. Poewe, C. Sampaio, G. T. Stebbins, M. B. Stern, B. C. Tilley, R. Dodel, B. Dubois et al., “Movement disorder society-sponsored revision of the unified parkinson’s disease rating scale (mds-updrs): process, format, and clinimetric testing plan,” Movement disorders, vol. 22, no. 1, pp. 41–47, 2007.

[6] C. G. Goetz, B. C. Tilley, S. R. Shaftman, G. T. Stebbins, S. Fahn, P. Martinez-Martin, W. Poewe, C. Sampaio, M. B. Stern, R. Dodel et al., “Movement disorder society-sponsored revision of the unified parkinson’s disease rating scale (mds-updrs): scale presentation and clinimetric testing results,” Movement disorders: official journal of the Movement Disorder Society, vol. 23, no. 15, pp. 2129–2170, 2008.

[7] P. Martinez-Martin, C. Rodriguez-Blazquez, M. Alvarez-Sanchez, T. Arakaki, A. Bergareche-Yarza, A. Chade, N. Garretto, O. Gershanik, M. M. Kurtis, J. C. Martinez-Castrillo et al., “Expanded and independent validation of the movement disorder society–unified parkinson’s disease rating scale (mds-updrs),” Journal of neurology, vol. 260, no. 1, pp. 228–236, 2013.

[8] G.-L. Defer, H. Widner, R.-M. Marié, P. Rémy, and M. Levivier, “Core assessment program for surgical interventional therapies in parkinson’s disease (capsit-pd),” Movement disorders: official journal of the Movement Disorder Society, vol. 14, no. 4, pp. 572–584, 1999.

[9] P. Martínez-Martín, A. Gil-Nagel, L. M. Gracia, J. B. Gómez, J. Martinez-Sarries, F. Bermejo, and C. M. Group, “Unified parkinson’s disease rating scale characteristics and structure,” Movement disorders, vol. 9, no. 1, pp. 76–83, 1994.

[10] G. T. Stebbins and C. G. Goetz, “Factor structure of the unified parkinson’s disease rating scale: motor examination section,” Movement disorders: official journal of the Movement Disorder Society, vol. 13, no. 4, pp. 633–636, 1998.

[11] G. T. Stebbins, C. G. Goetz, A. E. Lang, and E. Cubo, “Factor analysis of the motor section of the unified parkinson’s disease rating scale during the off-state,” Movement Disorders: Official Journal of the Movement Disorder Society, vol. 14, no. 4, pp. 585–589, 1999.

[12] J. Štochl, A. Boomsma, E. Tomešová, and K. Kovář, “Structural equation model of motor symptoms of parkinson’s disease,” Studies in Physical Culture & Tourism, vol. 13, 2006.

[13] S. D. Vassar, Y. M. Bordelon, R. D. Hays, N. Diaz, R. Rausch, C. Mao, and B. G. Vickrey, “Confirmatory factor analysis of the motor unified parkinson’s disease rating scale,” Parkinson’s Disease, vol. 2012, no. 1, p. 719167, 2012.

[14] E. J. Corti, A. R. Johnson, N. Gasson, R. S. Bucks, M. G. Thomas, and A. M. Loftus, “Factor structure of the ways of coping questionnaire in parkinson’s disease,” Parkinson’s Disease, vol. 2018, no. 1, p. 7128069, 2018.

[15] M. D. Humphries, J. A. Caballero, M. Evans, S. Maggi, and A. Singh, “Spectral estimation for detecting low-dimensional structure in networks using arbitrary null models,” Plos one, vol. 16, no. 7, p. e0254057, 2021.

[16] L. J. Evers, J. H. Krijthe, M. J. Meinders, B. R. Bloem, and T. M. Heskes, “Measuring parkinson’s disease over time: The real-world within-subject reliability of the mds-updrs,” Movement Disorders, vol. 34, no. 10, pp. 1480–1487, 2019.

[17] B. Harding, C. Tremblay, and D. Cousineau, “Standard errors: A review and evaluation of standard error estimators using monte carlo simulations,” The Quantitative Methods for Psychology, vol. 10, no. 2, pp. 107–123, 2014.

[18] S. Fahn, R. L. Elton, and U. P. Members, “Unified parkinson’s disease rating scale,” in Recent Developments in Parkinson’s Disease, S. Fahn, C. D. Marsden, M. Goldstein, and D. B. Calne, Eds. Florham Park, NJ: Macmillan Healthcare Information, 1987, vol. 2, pp. 153–163, 293–304.

[19] J. R. Evans, S. L. Mason, C. H. Williams-Gray, T. Foltynie, M. Trotter, and R. A. Barker, “The factor structure of the updrs as an index of disease progression in parkinson’s disease,” Journal of Parkinson’s disease, vol. 1, no. 1, pp. 75–82, 2011.

[20] C. R. Baumann, U. Held, P. O. Valko, M. Wienecke, and D. Waldvogel, “Body side and predominant motor features at the onset of parkinson’s disease are linked to motor and nonmotor progression,” Movement Disorders, vol. 29, no. 2, pp. 207–213, 2014.

[21] P. Riederer, K. Jellinger, P. Kolber, G. Hipp, J. Sian-Hülsmann, and R. Krüger, “Lateralisation in parkinson disease,” Cell and tissue research, vol. 373, no. 1, pp. 297–312, 2018.

[22] J. Seuthe, H. Hermanns, F. Hulzinga, N. D’Cruz, G. Deuschl, P. Ginis, A. Nieuwboer, and C. Schlenstedt, “Gait asymmetry and symptom laterality in parkinson’s disease: two of a kind?” Journal of Neurology, vol. 271, no. 7, pp. 4373–4382, 2024.

[23] M. H. Monje, Á. Śanchez-Ferro, J. A. Pineda-Pardo, L. Vela-Desojo, F. Alonso-Frech, and J. A. Obeso, “Motor onset topography and progression in parkinson’s disease: the upper limb is first,” Movement Disorders, vol. 36, no. 4, pp. 905–915, 2021.

[24] S. Zolfaghari, A. E. Thomann, N. Lewandowski, D. Trundell, F. Lipsmeier, G. Pagano, K. I. Taylor, and R. B. Postuma, “Self-report versus clinician examination in early parkinson’s disease,” Movement Disorders, vol. 37, no. 3, pp. 585–597, 2022.

[25] Kikuya, K. Tsukita, M. Sawamura, K. Yoshimura, and R. Takahashi, “Distinct clinical implications of patient-versus clinician-rated motor symptoms in parkinson’s disease,” Movement Disorders, vol. 39, no. 10, pp. 1799–1808, 2024.

[26] N. Hermanowicz, M. Castillo-Shell, A. McMean, J. Fishman, and J. D’Souza, “Patient and physician perceptions of disease management in parkinson’s disease: results from a us-based multicenter survey,” Neuropsychiatric Disease and Treatment, pp. 1487–1495, 2019.

[27] G. T. Stebbins, C. G. Goetz, D. J. Burn, J. Jankovic, T. K. Khoo, and B. C. Tilley, “How to identify tremor dominant and postural instability/gait difficulty groups with the movement disorder society unified parkinson’s disease rating scale: comparison with the unified parkinson’s disease rating scale,” Movement Disorders, vol. 28, no. 5, pp. 668–670, 2013.

[28] M. H. d. S. Tosin, C. G. Goetz, S. Luo, D. Choi, and G. T. Stebbins, “Item response theory analysis of the mds-updrs motor examination: tremor vs. nontremor items,” Movement Disorders, vol. 35, no. 9, pp. 1587–1595, 2020.

[29] B. A. Bernard, D. Carns, G. T. Stebbins, J. G. Goldman, and C. G. Goetz, “Relationship of movement disorders society–unified parkinson’s disease rating scale nonmotor symptoms to cognitive functioning in patients with parkinson’s disease,” Movement disorders clinical practice, vol. 7, no. 3, pp. 279–283, 2020.

[30] S. M. van Rooden, M. Visser, D. Verbaan, J. Marinus, and J. J. van Hilten, “Motor patterns in parkinson’s disease: a data-driven approach,” Movement disorders, vol. 24, no. 7, pp. 1042–1047, 2009.

[31] T. J. Delaney and C. O’Donnell, “Fast-local and slow-global neural ensembles in the mouse brain,” Network Neuroscience, vol. 7, no. 2, pp. 731–742, 2023.

[32] N. Rajamani, H. Friedrich, K. Butenko, T. Dembek, F. Lange, P. Navrátil, P. Zvarova, B. Hollunder, R. M. de Bie, V. J. Odekerken et al., “Deep brain stimulation of symptom-specific networks in parkinson’s disease,” Nature communications, vol. 15, no. 1, p. 4662, 2024.

